# Influence of Epigenetic Regulators Expression on Gastric Cancer Prognosis

**DOI:** 10.1101/2025.09.12.25335661

**Authors:** Lucas Brabo Rotella, André Salim Khayat, Monique Feitoza Silva, Victoria Pereira Costa, Diego Pereira, Fabiano Cordeiro Moreira, Samia Demachki, Samir Mansour Casseb, Geraldo Ishak, Williams Fernandes Barra, Rommel Mario Rodriguez Burbano, Paulo Pimentel de Assumpção

## Abstract

Gastric cancer (GC) is the fourth leading cause of cancer-related death worldwide. High mortality is largely associated with late diagnosis and tumor heterogeneity, highlighting the need for molecular insights. Epigenetic alterations, including histone modifications and DNA/RNA methylation, play a key role in gene regulation, tumor progression, and therapy response. This study analyzed 179 samples (156 tumor, 23 normal) from patients with gastric adenocarcinoma in Para, Brazil (2017 to 2020). RNA was extracted, quality checked, and used for cDNA library preparation (TruSeq, Illumina). Sequencing was performed on NextSeq 500, followed by quality control (FastQC), trimming (Trimmomatic), and quantification (Salmon, hg38/GENCODE v.42). Differential expression was assessed with DESeq2 in R. A panel of 304 epigenetic genes was examined, and expression patterns were visualized through heatmaps and clustering. Among the 304 genes analyzed, 167 (55%) showed differential expression in GC compared with normal tissue. Of these, 17 were related to DNA methylation, 29 to RNA methylation, 34 to histone acetylation, and 87 to histone methylation. Our results demonstrate dysregulation of all epigenetic processes investigated, promoting the activation of oncogenes (*DNMT1, MYC, ELF3, HDAC4,* and *HDAC7*) and the silencing of tumor suppressors (*CTCF, DNMT3B, ATRX, PRDM16* and *MSH6*), promoting genomic instability, apoptosis evasion, and tumor progression. No standard activation or repression patterns were found. Distinct and specific tumor profiles highlight the central role of epigenetic mechanisms in the pathogenesis of GC. Furthermore, epigenetic regulators have been shown to be potential biomarkers, prognosticators, and therapeutic targets.

## Introduction

Worldwide, Gastric Cancer (GC) is the fifth most common type and the fourth leading cause of cancer-related death [1]. In Brazil, the National Cancer Institute (INCA) estimated that between 2023 and 2025 there will be approximately 21,000 new cases of stomach cancer, ranking as the fourth leading cause of cancer mortality among men. In the Northern region of Brazil, gastric cancer is the second most common, with rates of 14.24 per 100,000 males and 7.57 per 100,000 females per year [2]. The occurrence in this region is influenced by a combination of environmental, infectious, and socioeconomic factors. Among these, regional dietary habits stand out, such as the high consumption of smoked and salted foods and the low intake of fruits and vegetables, which, together with late diagnosis, favor the emergence and persistence of epigenetic alterations [3].

High GC mortality rates are associated with late diagnosis and the lack of effective therapies that address the disease heterogeneity [4]. Despite grouping by stage and histological type, the diversity of prognoses and therapeutic responses reflects the limitations of current classification systems in capturing patient-specific variations. In this sense, epigenetic alterations play a critical role in gene expression regulation and, consequently, in histological phenotypic variations.

Epigenetics is characterized by reversible alteration in molecular structures that modulate gene expression without altering nucleotide sequences, through mechanisms such as histone modifications, DNA and RNA methylation, and microRNAs [5]. In cancer, epigenetics is fundamental in tumor progression, promoting silencing events in tumor suppressor genes and activating oncogenes. In addition, they may also contribute to the evasion of antitumor immunity, one of the hallmarks of cancer [6] by affecting tumor antigen presentation and effector immune cell functions [7, 8, 9].

In 2023, Weng et al. [10] proposed an epigenetic classification of gastric cancer, based on miRNA expression profiles and DNA methylation patterns obtained from databases (TCGA and GEO), which resulted in the definition of four molecular subtypes with distinct epigenetic profiles. These groups showed differences in prognosis and sensitivity to targeted therapies and chemotherapeutics such as 5-fluorouracil, paclitaxel, and cisplatin. Specific differences were also observed in cell proliferation pathways, tumor transformation, and ligand-receptor interactions, as well as distinctions in tumor stage distribution.

In this context, epigenetic studies attempt to clarify the mechanisms involved in cancer establishment and to identify new prognostic and diagnostic biomarkers, as well as potential candidates for targeted therapies. Therefore, this study aimed to investigate, in GC, differences in the expression of genes associated with the machinery of epigenetic mechanisms (histone methylation and acetylation, and DNA and RNA methylation), in order to understand the molecular factors of GC and contribute to the development of more precise and personalized therapeutic strategies.

## Materials and Methods

### Sample characterization and ethical considerations

In this study, 179 samples were analyzed: 156 from gastric tumor tissue and 23 from normal gastric tissue, obtained from patients diagnosed with gastric adenocarcinoma treated at the João de Barros Barreto University Hospital (HUJBB), Federal University of Pará (UFPA, Belém, PA, Brazil), and Ophir Loyola Hospital (HOL). Samples were collected between January 31, 2017, and July 29, 2020. All participants were informed about the objectives of the research, and samples were collected only after signing the Informed Consent Form. The study was approved by the Research Ethics Committee of HUJBB under protocol CAAE 10272913.8.0000.0017. Tumor fragments of 0.5 cm were collected immediately after gastric resection, stored in RNAlater for transport, and preserved at –80 °C.

### Total RNA extraction

Initially, tissue samples (50–100 ng) were homogenized and subjected to the addition of 1mL of TRIZOL® reagent for extraction, ensuring RNA integrity and cell lysis. After centrifugation at 13,000 rpm for 10 minutes at 4 °C, RNA was recovered by precipitation with isopropanol. The precipitated RNA was washed with ethanol, air-dried at room temperature, and quantified/assessed for integrity using a Qubit 2.0 Fluorometer and the 2200 TapeStation system. The following criteria were considered optimal: A260/A280 ratio between 1.8 and 2.2, A260/A230 ratio > 1.8, and RNA integrity number (RIN) ≥ 5. Total RNA was stored at –80 °C until further use.

### cDNA library construction

Libraries were prepared using the TruSeq Stranded Total RNA Library Prep kit with Ribo-Zero Gold (Illumina), following the manufacturer’s instructions. For each sample, 1μg of total RNA in a final volume of 10 μL was used. After construction, library integrity was reassessed using the 2200 TapeStation system.

### NGS sequencing

The cDNA libraries were loaded into the Illumina NextSeq system and subjected to paired-end sequencing (reads generated from both positive and negative cDNA strands). Sequencing was performed using the NextSeq® 500 ID Output V2 – 150 cycles kit (Illumina), following the manufacturer’s recommended conditions.

### Quality control and read alignment

Read quality was evaluated with FastQC. Adapters and low-quality sequences were removed using Trimmomatic v0.39. Human transcript reads were aligned and quantified using Salmon v1.5.2, with the reference index based on coding transcripts from hg38 (www.ensembl.org) and GENCODE v.42 annotation (www.gencodegenes.org). Reads were imported from Salmon into RStudio with the Tximport v3.14.0 package, and gene expression levels were estimated using the DESeq2 v3.14 package.

### Selection of genes to be investigated

A total of 304 genes related to the machinery of epigenetic mechanisms—histone methylation, histone acetylation, DNA and RNA methylation—were selected based on previous studies and databases such as WERAM. Selection prioritized genes with a direct impact on epigenetic processes, excluding those secondarily related to such processes and, in the case of RNA methylation, genes associated only with other RNA types rather than mRNA. All genes were categorized according to their respective epigenetic processes and identified by their impact on gene expression (associated with gene activation, repression, or variable impact).

### Statistical analysis

Differentially expressed genes were used to construct a heatmap in order to highlight global expression patterns between sample groups. For each gene, expression values were transformed into z-scores, allowing the standardization of relative variation and, consequently, direct comparison between samples. Visualization was performed using the ComplexHeatmap v2.25.2 package. Hierarchical clustering of samples were conducted using Spearman’s distance as the similarity metric and the average linkage method for dendrogram construction.

## Results

Of the 304 genes analyzed, 167 showed differential expression when comparing normal tissue versus cancer. Of these 167 differentially expressed genes, 74 exhibited a fold change of +2 /-2.

Differentially expressed genes related to DNA methylation machinery included 17 genes (Figure 1, top). The heatmap showed that GC samples exhibited a distinct expression profile compared to normal tissue. Regarding the gene classes in this process, there were 13 readers, 2 writers, and 2 erasers. When evaluating the impact on gene expression, 9 were associated with gene repression and 8 with activation. The genes with the highest fold change in this machinery were *SALL1* (FC=-9.73), *DNMT1* (FC=+2.85), *MYC* (FC=+2.74), *MBD1* (FC=+2.61), and *TET2* (FC=+2.57).

**Figure 1:**
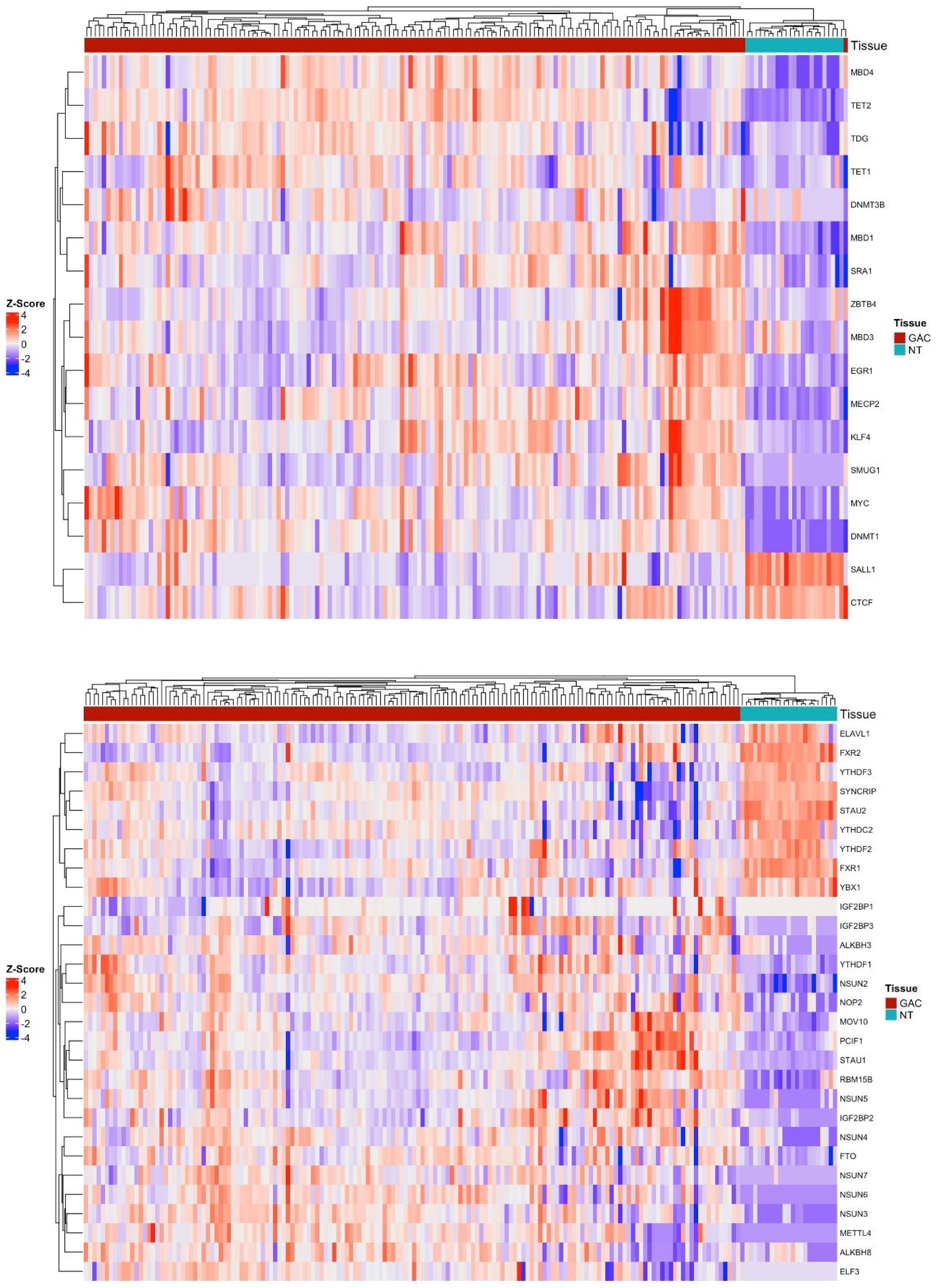
Top – Heatmap of DNA methylation machinery gene expression; Bottom – Heatmap of RNA methylation machinery gene expression.

Regarding RNA methylation, 29 genes were differentially expressed (Figure 1, bottom). In this machinery, a difference in gene expression profiles was also observed, with a notable number of reductions in the activity of genes that are normally active. The classification of results included 12 readers, 10 writers, 3 erasers, 3 transcription factors, and 1 regulator. For RNA methylation, it is not viable to classify the impact as activation or repression. The genes with the highest fold change in this machinery were *IGF2BP1* (FC=+15.5), *FXR2* (FC=-3.57), *ELF3* (FC=+3.15), *METTL4* (FC=+3.02), and *STAU* (FC=-2.72).

The results of histone modifications, starting with the acetylation process, showed a total of 34 differentially expressed genes, including 13 readers, 13 histone deacetylases, 7 histone acetyltransferases, and 1 transcription factor. The heatmap (Figure 2, top) of these genes demonstrates inversions in expression activity for some genes, with a prominent cluster of GC samples showing low expression of genes normally active and high expression of genes normally lowly expressed. Sixteen genes were associated with activation, 14 with repression, and 4 showed context-dependent effects. The genes with the highest fold change in the acetylation process were DPF1 (FC=-9.08), TAF1L (FC=-7.65), HDAC10 (FC=+3.83), DPF3 (FC=-3.78), and MLLT1 (FC=+3.59).

**Figure 2:**
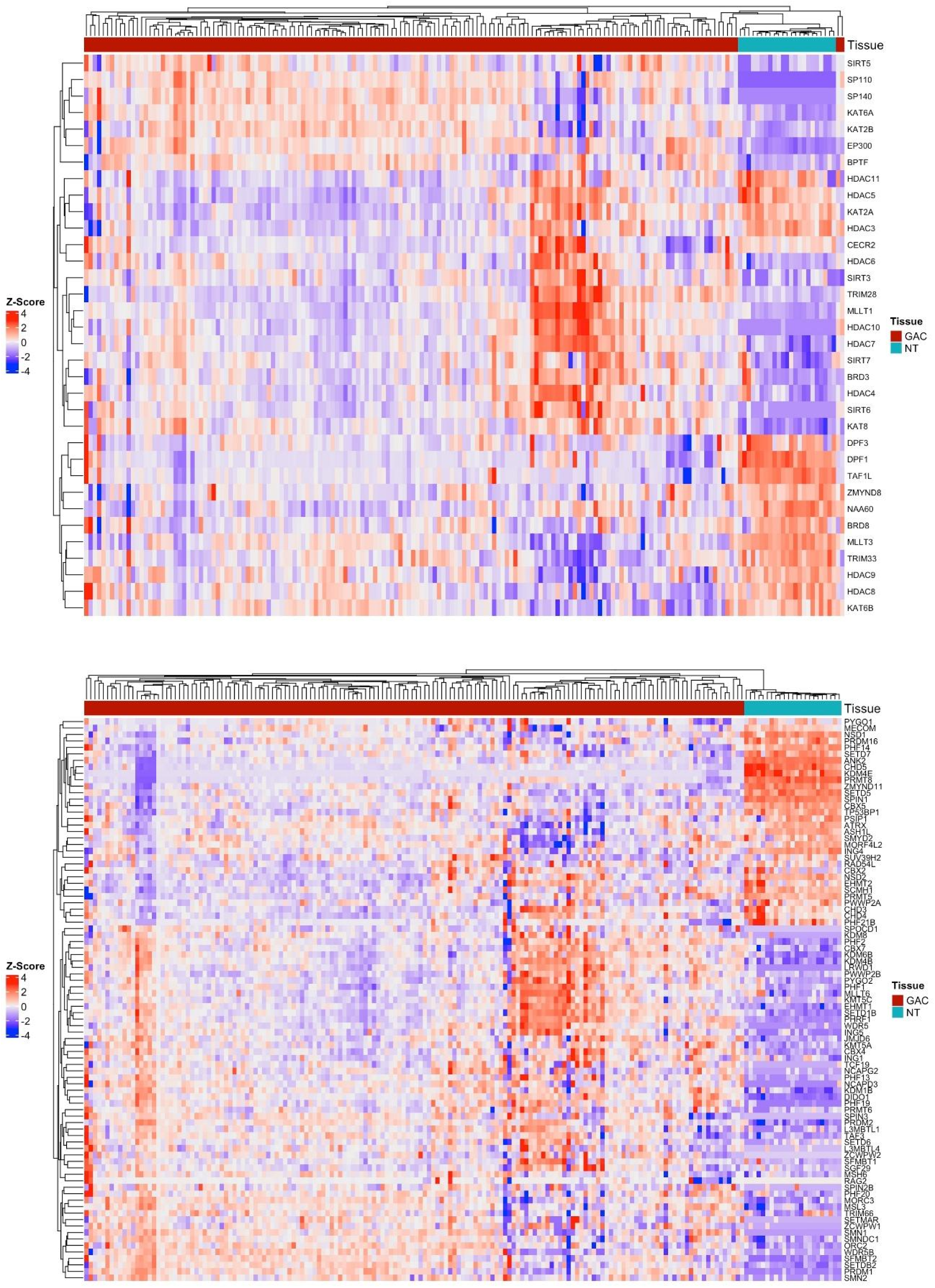
Top – Heatmap of histone acetylation machinery gene expression; Bottom – Heatmap of histone methylation machinery gene expression.

Finally, the results of the histone methylation machinery included 87 differentially expressed genes: 58 readers, 22 histone methyltransferases, and 7 histone demethylases. Of these, 35 were associated with repression, 24 with gene activation, and 28 showed context-dependent effects. The heatmap of these genes (Figure 2, bottom) highlights, among the observed variations, a pattern of low expression in all GC samples for genes normally highly expressed in normal tissue. The genes with the highest fold change in this epigenetic process were *PRMT8* (FC=+11.11), *RAG2* (FC=+10.67), *ANK2* (FC=-10.65), *CHD5* (FC=-10.24), and *KDM4E* (FC=-7.59).

Grouping genes by their impact on gene expression, there were 47 differentially expressed genes related to activation (DNA demethylation, histone acetylation, and histone methylation/demethylation) and 59 related to repression (DNA methylation, histone demethylation, and histone methylation/demethylation). For this grouping, 32 genes with variable impact and 29 genes associated with RNA methylation were excluded. For both repression and activation, the most frequent epigenetic processes were histone methylation/demethylation, histone acetylation/deacetylation, and DNA methylation/demethylation, respectively.

The genes with the highest fold change in the heatmap of expression-related genes (Figure 3, top) were *RAG2* (FC=+10.67), *DPF1* (FC=-9.09), *TAF1L* (FC=-7.65), *KDM4E* (FC=-7.59), and *PRDM16* (FC=-7.57). In the heatmap of repression-related genes (Figure 3, bottom), the genes with the highest fold change were *CHD5* (FC=-10.24), *SALL1* (FC=-9.73), *PHF21B* (FC=-7.46), *HDAC10* (FC=+3.83), and *SIRT6* (FC=+3.33).

**Figure 3:**
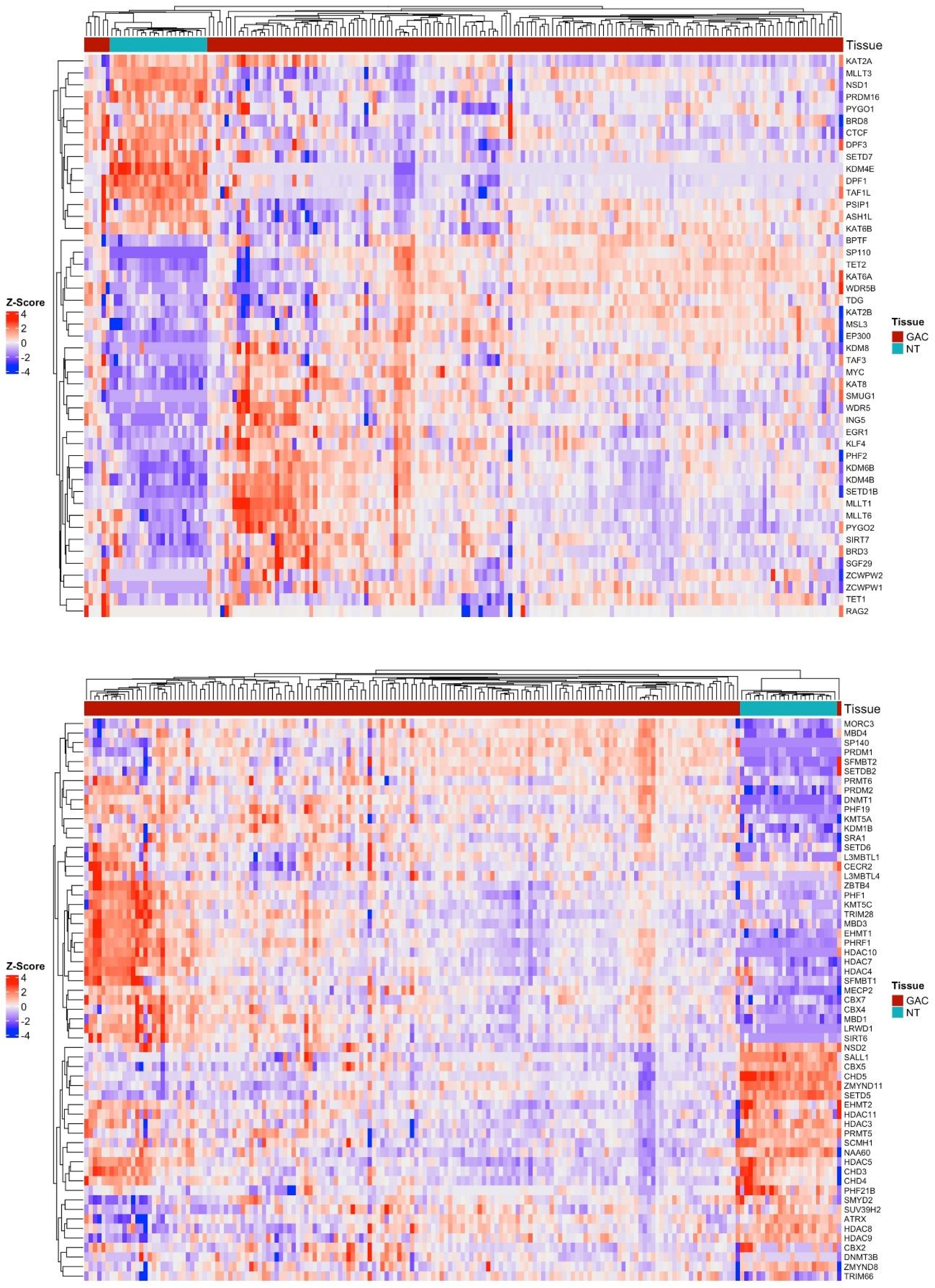
Top – Heatmap of genes related to gene expression activation; Bottom – Heatmap of genes related to gene expression repression.

## Discussion

Unlike genetic mutations, epigenetic modifications dynamically and reversibly alter gene expression, modulating the activity of genes important for carcinogenesis, such as oncogenes and tumor suppressors. Among the main epigenetic processes, DNA methylation, RNA methylation, and histone modifications stand out, all of which were found to be altered in our results in gastric cancer samples compared to normal tissue. Of the genes analyzed, 55% were found to be altered, including 13 oncogenes—*CHD4, MECOM, NSD1, NSD2, PHF19, BRD3, HDAC4, HDAC7, MLLT3, ELF3, DNMT1, KLF4,* and *MYC—*and 18 tumor suppressors—*CTCF, DNMT3, BEGR1, KLF4, MBD4, TET1, TET2, ELF3, EP300, HDAC4, SP140, ATRX, ING1, MSH6, NSD1, PRDM1, SETDB2,* and *TP53BP1*—which were analyzed within their respective epigenetic machinery contexts according to their hyper- or hypoexpression.

Tumor suppressor expression is often reduced in cancers due to inactivation mechanisms, including epigenetic silencing, which confers a selective advantage for uncontrolled cell proliferation [11]. The presence of upregulated tumor suppressors in our study (*EGR1, TET1, TET2, EP300, SP140, HDAC4, ING1, MSH6, PRDM1,* and *SETDB2*) may be associated with a response to oncogenic stress, which is, through mechanisms essential for tumor survival, neutralized by other molecular alterations without the need to silence the gene.

The first machinery analyzed, DNA methylation, constitutes an epigenetic process with diverse impacts on carcinogenesis. Aberrant DNA methylation in cancer can be divided into hypomethylation and hypermethylation. Hypomethylation is associated with genomic instability, cancer progression, loss of imprinting, and oncogene activation. On the other hand, hypermethylation is a major mechanism for inactivating tumor suppressor genes, as well as affecting cell cycle control, DNA repair, cell adhesion, invasion, migration, and apoptosis [12, 13].

Our results highlight several alterations in the expression of genes associated with the DNA methylation/demethylation process. In GC samples, overexpression of genes normally underexpressed and the underexpression of genes normally overexpressed can be observed. This finding not only corroborates the correlation between aberrant methylation and cancer but also favors the occurrence of aberrant methylation itself by altering the expression of genes responsible for controlling this process.

Among the differentially expressed genes, the overexpression of oncogenes *DNMT1* (FC=+2.85)—associated with resistance to 5-Azacitidine treatment, being a potential biomarker, correlated with a poorer prognosis [14, 15]—and *MYC* (FC=+2.74)—linked to aggressive GC, poor prognosis, and involved in cell proliferation, angiogenesis, immortalization, apoptosis evasion, and metastasis [16, 17]—stands out. Regarding underexpressed tumor suppressor genes, *CTCF* (FC=-1.01) and *DNMT3B* (FC=-1.40) were observed, both associated with genomic instability and silencing of other tumor suppressors [18, 19]. Other relevant genes in our results include *KLF4, SALL1*, and *MBD1*, although their impacts on GC are lower described or less significant compared to the previously mentioned genes.

The second epigenetic process, RNA methylation, involves N6-methyladenosine (m6A) methylation, participates in RNA splicing, and also affects protein synthesis by interfering with mRNA stability and, consequently, the duration of synthesized mRNA available for translation [20]. This process has also been reported to be associated with the initiation and development of cancers, including glioma, esophageal, colorectal, and gastric cancer [21]. Biological functions affected by altered RNA methylation include mRNA export, degradation, and translation. In the context of carcinogenesis, it is involved in proliferation, migration, tumor invasion, and aberrant cell differentiation [22, 2

Analysis of this machinery revealed an aberrant pattern compared to the nearly identical pattern observed in normal samples, reinforcing the association of epigenetic modifications with cancer emergence and progression. RNA methylation is still in the early stages of investigation in various cancers, however, several genes with described impacts can be mentioned, such as the oncogene *ELF3* (FC=+3.15), overexpressed in our results and associated with poorer prognosis [24], along with *IGF2BP1* (FC=+15.48)—promoting cancer progression and exacerbated mRNA stabilization and translation [25, 26]—and *METTL4* (FC=+3.02)—promoting metastasis, migration, and tumor invasion [27, 28], both also overexpressed.

Regarding histone modifications, acetylation and methylation direct the accessibility of cellular genetic information, allowing or inhibiting processes such as transcription, replication, and DNA repair—key events in cancer progression [29]. Dysregulation of histone modifications, including aberrant methylation and acetylation, is associated with GC, tumor progression, growth, and metastasis. Altered methylation correlates with the positive regulation of cell–cell and cell–extracellular matrix adhesion genes, facilitating invasion and, consequently, poorer prognosis [30]. Aberrant acetylation promotes tumor growth, chemoresistance, and metastasis, while also favoring the silencing of tumor suppressor genes [31].

The results of both histone modification processes also highlight expression profiles in GC that are distinct from normal tissue, corroborating, as with DNA methylation, the associations already described in the literature. Therefore, the importance of alterations in this machinery for carcinogenesis is reaffirmed, representing a critical step that enables the modulation of other genes, such as oncogenes and tumor suppressors, in a manner that promotes tumor emergence and progression.

In the histone acetylation machinery, overexpression of *MLLT1* (FC=+3.59), *HDAC10* (FC=+3.82), and *SIRT6* (FC=+3.33) stands out; these genes are directly associated with tumor progression and poorer clinical prognosis in various cancer types [32, 33, 34], reinforcing the impact of epigenetic reprogramming on GC aggressiveness. Overexpression of *BRD3*, a member of the BET family, was also observed; as a reader, it recognizes acetylated histones and promotes transcriptional activation of oncogenic programs [35]. Additionally, the overexpressed oncogenes *HDAC4* (FC=+1.33) and *HDAC7* (FC=+2.44) – class IIa deacetylases acting as erasers – remove acetylation marks, leading to chromatin repression and participating in the silencing of tumor suppressor genes, thereby favoring tumor plasticity and apoptotic evasion [36].

This imbalance between activation (via BET) and repression (via HDACs) of histone acetylation creates a transcriptional instability landscape that favors the development of therapeutic resistance, emphasizing the relevance of epigenetic approaches as therapeutic targets in gastric cancer, both through *HDAC* inhibition [37] and BET activity blockade [38].

When analyzing histone methylation results, the *CBX7* gene—a component of the Polycomb repressive complex—stands out for exhibiting overexpression (FC=+2,20), as its activity is associated with the modulation of fundamental cellular processes such as proliferation and migration. Recent hypotheses propose that this gene regulates the gastric tumor stem cell niche, thereby influencing tumorigenesis and disease progression [39].

In addition, within this machinery, our gene expression analyses revealed notable underexpression of important genes such as *CHD5* (FC=–10.24) and *PRDM16* (FC=-7.57). Such negative regulation is generally mediated by hypermethylation of their promoters, an epigenetic event that silences their transcription and function. Although they act in distinct pathways, the convergent alteration of these genes indicates aggressiveness and rapid progression of gastric carcinogenesis.

The *CHD5* gene is a chromatin remodeler that, when altered, disrupts cell cycle control and promotes genomic instability. This gene represents an interesting biomarker, as previous analyses have associated it with lymph node metastasis and poorer prognosis [40]. In addition, analysis of the *PRDM16* gene is of particular interest due to its contrasting impact when mutated in hematologic cancers versus solid tumors, raising the possibility of therapeutic targets in gastric cancer. As a histone methyltransferase, alterations in this gene may lead to loss of cellular identity through its association with differentiation and proliferation events in GC, particularly in relation to stem cell regulation [41].

Although to a lesser extent, we also observed the underexpression of an important tumor suppressor gene. *ATRX* (FC=–1.44) plays an important role in the control of cellular senescence. Its activity is linked to alternative lengthening of telomeres, and when mutated, it contributes to replicative immortality and aggressive tumor growth [42].

All epigenetic processes were initially analyzed together and, subsequently, genes were grouped according to their impact on activation or repression of gene expression. The aim was to investigate whether there was a predominance of global patterns of chromatin decondensation or condensation, reflecting a general trend toward increased expression or repression in gastric cancer compared to normal tissue.

Although the results obtained did not reveal a consolidated pattern in GC for this evaluation, both gene groups (activation and repression) exhibited expression profiles distinct from normal tissue, highlighting that alterations in both pathways are necessary for carcinogenesis. Beyond the previously discussed targets (oncogenes and tumor suppressors), the changes observed across all epigenetic processes in this study indicate that various alterations in the expression of known genes in GC may be potentially associated with cancer’s control over epigenetic regulation, and could, in fact, be a consequence of such control.

However, no consolidated pattern in GC was observed. Nonetheless, both the gene groups associated with activation and repression showed distinct profiles compared to normal tissue, highlighting the relevance of dysregulation of both processes in GC. In addition to the oncogenes and tumor suppressors already discussed, the alterations identified across all epigenetic processes raise the hypothesis that many previously described changes in gene expression in GC may be potential consequences of cancer’s control over epigenetic regulation.

## Conclusion

Moreover, our results demonstrated that the main epigenetic processes are broadly dysregulated in gastric cancer, promoting both the activation of oncogenes (*DNMT1, MYC, ELF3, HDAC4,* and *HDAC7*) and the silencing of tumor suppressors (*CTCF, DNMT3B, ATRX, PRDM16* and *MSH6*), creating a scenario of genomic instability, apoptosis evasion, and tumor progression. Although no overarching pattern of greater activation or repression was identified, the distinct profiles observed in tumor tissue reinforce the importance of alterations in both processes for GC, highlighting epigenetic regulation as a key mechanism in the disease and suggesting that genes within the epigenetic machinery could act as potential prognostic biomarkers and therapeutic targets.

## Data Availability

All data produced in the present study are available upon reasonable request to the authors

## Acknowledgment

The authors would like to thank the Oncology Research Center and the Human and Medical Genetics Laboratory at the Federal University of Pará (UFPA) for their invaluable technical and laboratory support. We are also grateful to the High-Performance Computing Center (CCAD) at UFPA for providing access to the Apollo 2000 cluster, which was essential for our analyses.

